# Deep Learning for Predicting Cognitive Gap as a Reliable Biomarker of Dementia

**DOI:** 10.1101/2021.01.24.21249625

**Authors:** Tetiana Habuza, Nazar Zaki, Yauhen Statsenko, Fady Alnajjar, Sanaa Elyassami

## Abstract

Neuroimaging data may reflect the mental status of both cognitively preserved individuals and patients with neurodegenerative diseases. To find the relationship between cognitive performance and the difference between predicted and observed functional test results, we developed a Convolutional Neural Network (CNN) based regression model to estimate the level of cognitive decline from preprocessed T1-weighted MRI images. In this study, we considered the Predicted Cognitive Gap (PCG) as the biomarker to accurately classify Healthy Control (HC) subjects versus Alzheimer disease (AD) subjects. The proposed model was tested on a dataset that includes 422 HC and 377 AD cases. The performance of the proposed solution was measured using Receiver Operating Characteristic (ROC) Area Under the Curve (AUC) and achieved 0.987 (ADAS-cog), 0.978 (MMSE), 0.898 (RAVLT), 0.848 (TMT), 0.829 (DSST) for averaged brain images; and 0.985 (ADAS-cog), 0.987 (MMSE), 0.901 (RAVLT), 0.8474 (TMT), 0.796 (DSST) for middle slice skull stripped brain images. The results achieved indicate that PCG can accurately separate healthy subjects from demented ones and thus, the structure of the brain contributes to the level of human cognition and their functional abilities. Therefore, PCG could be used as a biomarker for dementia.

## I. Introduction

Brain atrophy is an essential subject of dementia-related research. It’s a condition in which the brain or specific regions of the brain shrink in size. Currently, 50 million people worldwide are unfortunately living with dementia, and this number is expected to double every 20 years to reach over 130 million by 2050 [1]. Aging of the society is a typical problem in developed countries, which leads to high incidences of dementia and other Neurodegenerative Diseases (NDs). Several neuropsychological test batteries can objectively and reliably assess a subject’s cognitive abilities. The cognitive and functional assessment is a proven approach to diagnose irreversible, progressive brain disorders, such as Alzheimer’s Disease (AD), that destroys thinking and memory skills. However, it requires creating a productive testing environment to keep the subject focused along with managing the testing time. Usually, the procedure of assessment of the subject’s cognitive abilities is time-consuming.

Biomarkers may be used to distinguish different aspects of pathology; detect pre-symptomatic pathological changes; predict decline or conversion between clinical disease states; or monitor disease progression and response to treatment [2]. Figure 1 illustrates the most common biomarkers that can identify dementia in different settings.

**Figure 1:**
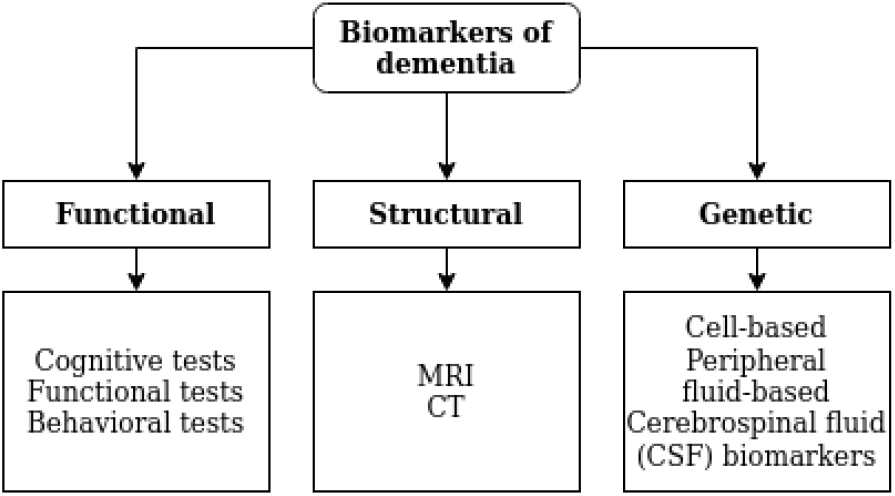
Different modalities of dementia’s biomarkers

Although several previous studies investigated the structural changes or the functional impairment of the nervous system (e.g., cognitive decline), it is still unclear, to what degree brain atrophy contributes to the dysfunction of the nervous system. Structural magnetic resonance imaging (MRI) studies have revealed that the extent of age-related brain changes varies markedly across individuals [3]. Other studies of brain functioning bared inconsistencies in both onset and the rate of episodic memory loss in the elderly cohort, which accounts for different inherited and lifestyle factors. However, there is no evidence of a direct link between structural and functional impairment.

Fukurama et al. [4] emphasized the importance to understand the relationship between functional age-related change and neurological dysfunction. In this study, the authors focused on cerebral- and neuro-imaging, which are a range of tools used to visualize the structure, functions, and pathogenic molecules in the nervous system. This is in line with the recent study by Vernooij et al. [5], where through a European-wide survey, the authors assessed the current clinical practice of imaging in the primary evaluation of dementia, to standardized imaging, evaluation, and reporting. The authors showed the importance of using visual rating scales, implementation of volumetric assessment, and structured reporting to improve the diagnosis.

Published studies on existing dementia risk models most commonly comprised from demographics, subjective cognitive complaints, cognitive test scores, lifestyle and health-related variables [6]. Several studies focused on predicting the risk of dementia and used cognitive test scores or neuropsychological test batteries as predictors. The most commonly used cognitive variables are Mini-Mental State Examination (MMSE), Rey Auditory-Verbal Learning Test (RAVLT), Alzheimer’s Disease Assessment Scale cognitive subscale (ADAS), Digit Symbol Substitution Test (DSST), Logical Memory Delayed Recall Test, Trail Making Test [6]. Although MMSE is often used, it shows only moderate predictive accuracy [7] in risk models, whereas RAVLT shows stronger predictive potential and association with dementia [8]. In [9] the authors illustrated the boost in the predictive performance of the diagnosis when cognitive features were ensemble with structural features. Ultimately, if a correlation between imaging data and the cognitive test results exists, then we can use it as a reliable marker.

As machine learning (ML) and Deep Learning (DL) algorithms continue to show reliable performances, several authors have started to apply them to brain atrophy research. The authors in [10] provided detailed overview of the most recent research in dementia diagnosis using biomedical image processed using ML techniques. The authors highlighted the state-of-the-art approaches in neuroimaging, brain anatomy assessment and ND diagnosis. The authors then concluded that the broader use of neuroimaging will lead to the accumulation of large datasets which will improve the overall diagnostic capabilities. In spite of the promising ability to diagnose AD using MRI scans, substantial progress is still needed [11], [12].

In addition, recent studies demonstrated that the extent of the microstructural changes in the limbic system on brain MRI images correlates with the magnitude of cognitive impairment [13]. Brain atrophy assessed on structural MRI shown a valid marker of Alzheimer’s disease (AD) [14] and AD-related neurodegeneration at the late stages of the disease. However, reliable means of identifying cognitively normal individuals at higher risk to develop AD are more likely to derive from psychophysiological testing (e.g., event-related potentials) [6], [15]. Ultimately, the full understanding of the pathophysiological mechanisms underlying AD-and mild cognitive impairment (MCI)-related functional impairment of the brain and its structural bases remains incomplete [4]. Therefore, studying brain atrophy and dementia is very attractive for researchers nowadays as new findings may have a huge impact on earlier detection, diagnosis, and treatment of age-related degenerative diseases.

In this paper, we provide a comparative analysis of different cognitive tests predicted from structural brain features by evaluating the PCG of the healthy and nonhealthy controls from their T1 weighted MRI images. Our objective is to find the possible relationship between cognitive performance and PCG.

To address the objective of the study we formulate the following tasks

- Conduct the exploratory analysis of the functional features for the study cohorts with cognitively-preserved people and patients with Dementia (AD)
- Propose the methodology of predicting the PCG out of T1-weighted images of the brain
- Justify the PCG as a reliable biomarker of disease-related cognitive impairment

## Methods

### A. Dataset

The data used in this study was obtained from the Alzheimer’s disease neuroimaging initiative (ADNI) database (www.adni-info.org). From ADNI1 dataset [16] we acquired clinical information on 799 cases (HC/AD 52/82%/47.18%); male/female 54.07%/45.93%), along with cognitive assessment results, demographic and morphometric features.

### B. Cognitive and functional assessments

To assess a subject cognitive ability, all ADNI subjects underwent standardized neuropsychological testing which include the following:

- **Mini Mental state examination (MMSE):** The most commonly used test for complaints of problems for memory or other mental abilities. It consists of a questionnaire that is used to measure cognitive impairment. It is also used in the estimation of the severity and progression of cognitive impairment [17]. The maximum MMSE score is 30 points, normal range (from 25 to 30), mild dementia (from 20 to 24), moderate dementia (from 13 to 19), and severe dementia (less or equal to 12). On average, the MMSE score of a person with AD declines about two to four points a year.
- **Rey Auditory-Verbal Learning Test (RAVLT):** Designed to evaluate the episodic and verbal memory in patients. The RAVLT assesses the ability to acquire 15 words across five immediate learning trials, to recall the words immediately after an intervening interference list, and to recall and recognize the words after a 30-minute delay. This test can be used to evaluate the level of memory dysfunction and track it over time [18].
- **Trail Making Test (TMT):** Evaluates processing speed and executive function. It consists of two parts. Both Parts A and B depend on visual-motor and perceptual-scanning skills, but Part B also requires cognitive flexibility in shifting from number to letter sets under time pressure [19]. In ADNI part B is used, which calculates the total number of seconds to complete Part B, up to a maximum of 300 seconds.
- **Digit Symbol Substitution Test (DSST):** It consists of some number (e.g. nine) of digit-symbol pairs (e.g. 1/-,2/+ …7/ ,8/x,9/=) followed by a list of digits. Under each digit, the subject should write down the corresponding symbol as fast as possible and the number of correct symbols within the allowed time (e.g. 90 or 120 sec) is measured [20].
- **Alzheimer’s disease assessment scale – cognitive (ADAS-cog):** A brief cognitive test battery that assesses learning and memory, language production, language comprehension, constructional praxis. It assesses the subject’s ability to copy four geometric figures ranging from a very simple one to a difficult one), ideational praxis and orientation [21].

### C. Data pre-processing

The pre-processed T1 weighted sMRI images were downloaded in NIFTI format along with clinical data from [16].

To enhance the predictive performance metrics, the nonbrain tissues were extracted from the image using BET (Brain Extraction Tool) [22]. It deletes non-brain tissue from an image of the whole head [23]. Following the brain extraction and to reduce the computational complexity, we calculated the average of the voxel intensities values over the z-axis in the axial view of the extracted brain to obtain a two-dimensional brain image. Besides, we extracted only one slice of the brain from the middle of the axis Z in the axial view. Then the data were down-sampled with nearest-neighbor interpolation to the size of 150 by 150 pixels, normalized within the range from 0 to 1, and stored in JPEG format. A sample extracted image is shown in Figure 2.

**Figure 2:**
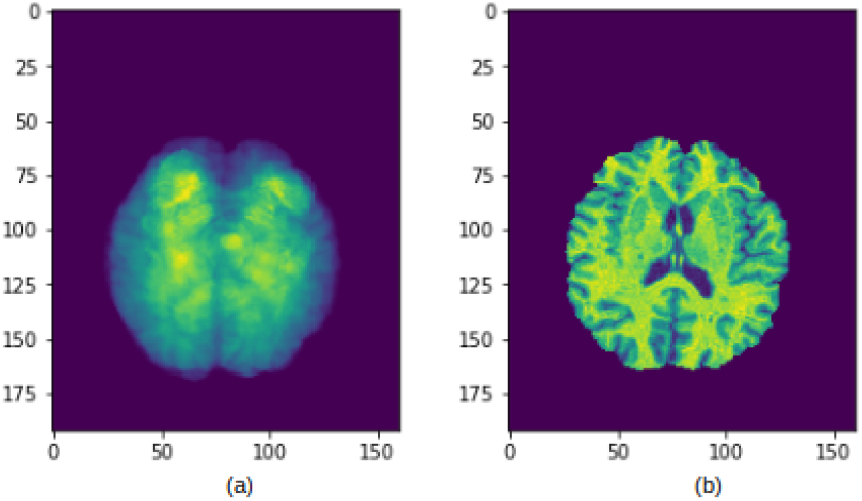
A sample extracted image following the proposed pre-processing steps (a) Averaged over the z-axis in axial view brain, extracted with BET tool; (b) The middle slice of the skull-stripped brain obtained in axial view.

For the unification of the pre-processing workflow, we used Nipype, an open-source, community-developed initiative under NiPy [24]. To automate the deployment of applications inside software containers we used Neurodocker that wraps up the aforementioned software in a complete package.

### D. Proposed design and methods

The workflow of the proposed method is illustrated in Figure 3.

**Figure 3:**
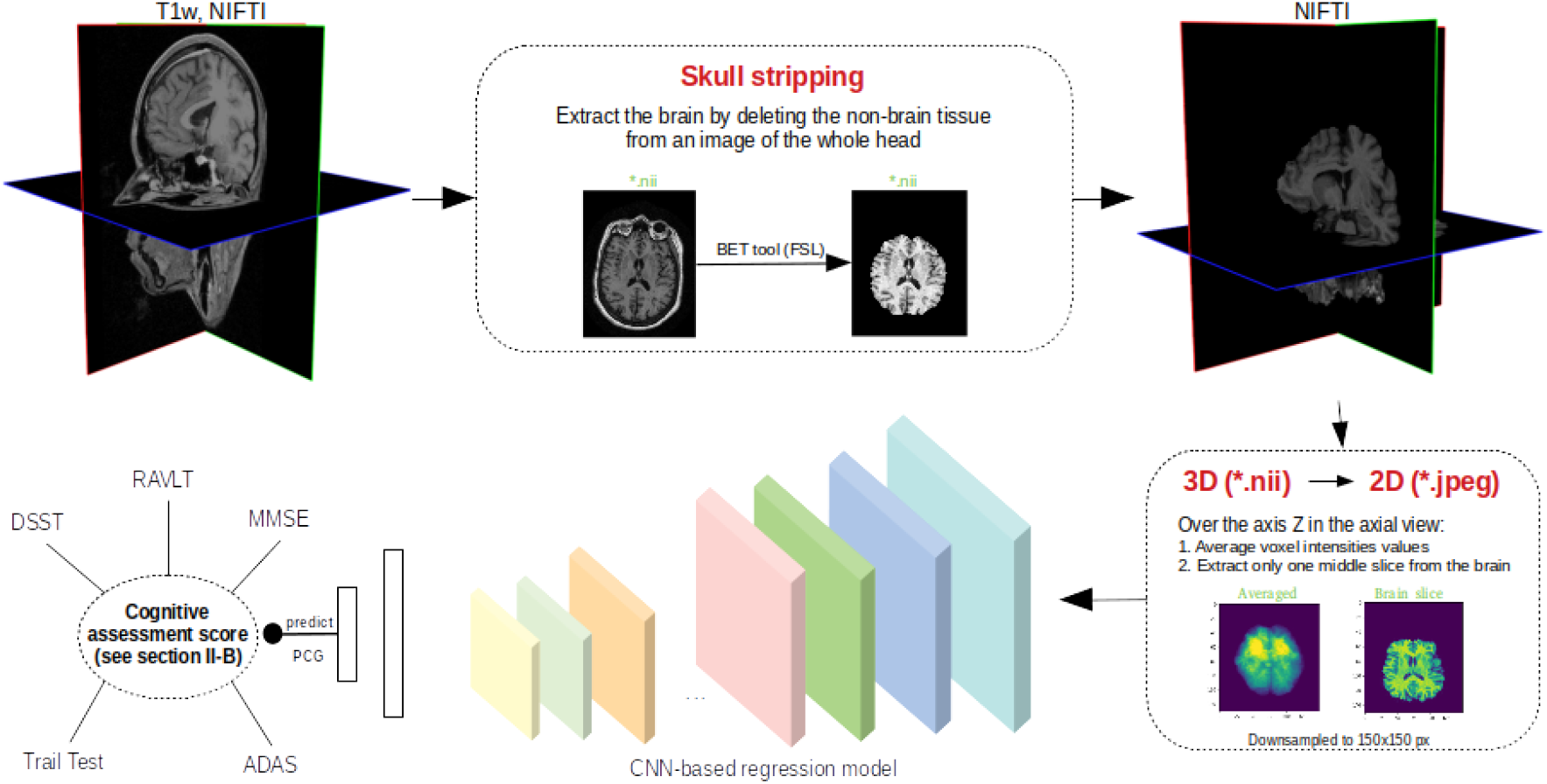
The proposed pipeline of finding reliable functional assessment of the cognitive status using MRI structural biomarkers.

First, we evaluated the separability of the two different disease groups utilizing non-parametric statistic methods. Mann-Whitney U test was used for the continuous features and Fisher’s Exact Test for the quantitative ones. In this case, the data was expressed as *IQR, mean* ± *std* or number of cases and their percentage.

Second, we assessed the reliability of the functional assessments using structural MRI biomarkers by evaluating the gap between predicted and actual values of the cognitive test results for healthy controls and people with dementia. We developed a DL model to predict the cognitive gap. Over the past decade, DL techniques have shown an enormous breakthrough in the field of medical diagnostics by achieving excellent performance in automatic medical imaging classification, detection, and segmentation [25].

The architecture of the proposed CNN regression model is similar to AlexNet. It consists of six convolution layers followed by two fully connected dense layers. The model was also regularized using L2 penalty with 0.0001 alpha value. We used RMSProp optimizer and trained the network for 100 epochs or before converged using. By convergence, we mean that the validation loss function is not improving by at least 0.0001 for 10 consecutive iterations. We used 20% of training data for validation purposes. The model was trained using a five-fold cross-validation technique on a healthy population and tested on two disease cohorts such as HC and AD. The outcome of the model is a set of PCG values calculated for HC and AD cohorts.

Finally, following the learning step, we performed a comparative analysis of the PCG for healthy and dmented populations. We calculated the mean and standard deviation for the PCG values in two cohorts and compared their distributions. Next, we build another machine learning classification model to predict disease using PCG values. To measure the accuracy of the performance, we calculated the sensitivity and sensitivity of PCG for the MMSE, ADAS-dog, TMT(part B), RAVLT, and DSST tests. An accurate prediction can be used as a potential early biomarker of neurodegeneration and a way for early intervention.

This research work was implemented mainly in Python, and it’s supporting libraries for Computer Vision, Data Visualization, Data Processing, such as NumPy, Pandas, SciPy, Matplotlib, PIL, Pillow, OpenCV, Scikit-learn. We also used neurodocker container with functioning Python, Nipype, FSL, ANTs, and SPM12 software packages in addition to a TensorFlow-GPU-based container. Finally, the experimental work was run on Linux Ubuntu 18.04 Nvidia DGX-1 deep learning server accessed with web-based multi-user concurrent job scheduling system [26].

## III. Experimental work and results

### A. Comparison of the cognitively preserved vs demented population

The comparative analysis of the subjects with regards to age, gender, and the cognitive test results are presented in Table I. It shows no significant difference between HC and AD groups concerning age and gender, so our predictions will fairly interpret the disease outcome.

**Table I:** Comparison of subjects with regards to the cognitive and structural features outcomes

|  | Total<br>n=799 | CN<br>422(52.82%) | Dementia<br>377(47.18%) | p-value |
| --- | --- | --- | --- | --- |
| AGE | 75.7[71.6-80.1] | 75.99±5.37 | 75.38±7.34 | 0.3038 |
| Gender |  |  |  | 0.7223 |
| Male | 432(54.07%) | 231(54.74%) | 201(53.32%) |  |
| Female | 367(45.93%) | 191(45.26%) | 176(46.68%) |  |
| Cognitive tests |  |  |  |  |
| CDR | 2.32[0.0-4.5] | 0.06±0.21 | 4.86±2.1 | p<0.01 |
| ADAS-cog | 37.25[16.0-56.66] | 17.83±8.41 | 58.98±17.03 | p<0.01 |
| MMSE | 26.07[23.0-29.0] | 29.11±1.05 | 22.66±3.03 | p<0.01 |
| RAVLT | 40.45[28.0-53.0] | 51.73±10.76 | 27.82±9.87 | p<0.01 |
| DSST | 36.9[26.5-47.0] | 46.62±10.47 | 26.02±12.31 | p<0.01 |
| TMT(part B) | 136.98[69.0-190.25] | 83.87±39.56 | 201.41±88.32 | p<0.01 |

*Functional variables* consist of some cognitive test results conducted in ADNI1 study (see section II-B). ADAS-cog (*ADAS* = *ADAS*_4_ + *ADAS*_11_ + *ADAS*_13_) and RAVLT (*RAV LT* = *RAV LT*_*immediate*_ + *RAV LT*_*learning*_ + *RAV LT*_*f orgetting*_) in the table correspond to the cumulative values of corresponding tests (for more details see [16]). There are significant differences between disease groups with regard to the cognitive assessment. Our subjects are clearly separable by MMSE, ADAS-cog, RAVLT, TMT(part B), and DSST functional outcomes.

### B. Predicted Cognitive Gap (PCG)

We denote PCG as a difference between the outcome predicted by our model and actual cognitive test result and assume that it can serve as a biomarker to screen for dementia. It is consistent with studies related to brain age gap biomarker [27], [28].

To predict the level of cognitive decline preprocessed image data were fed to the designed regression CNN-based model. Target or dependent variables were chosen from the results of the cognitive assessment such as MMSE, ADAS-cog, RAVLT, TMT(part B), and DSST.

The CNN-based model was trained in a five-fold cross-validation manner on the averaged and middle slice brain images obtained from a healthy population and then tested on two different groups such as HC and AD. The performance of our model for all five cognitive tests can be found in Table II. We estimate the quality of the model in terms of Mean Absolute Error (MAE) and highlight it in bold.

**Table II:**
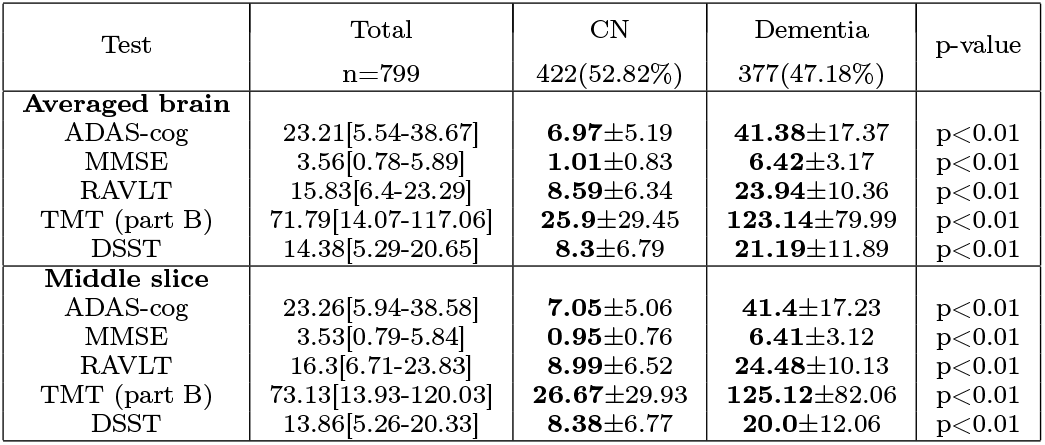
Comparison of predicted cognitive gap in the healthy population and population with dementia

From Table II, there is a notable difference between the performance of the models. MMSE was predicted more accurately from middle brain-slice data, whereas averaged MRI brain images were the predictors of choice for forecasting ADAS-cog, DSST, TMT, and RAVLT. In general, the better performance was obtained on the averaged data. The possible explanation for this is that a middle slice misses full information on the structures that are not trapped in it. In contrast to this, an averaged image somehow accumulates the data for the entire brain.

### C. Justification of PCG as a biomarker of dementia

First, to justify the PCG as a reliable biomarker of dementia, we conducted a comparative analysis of PCG for different disease cohorts (Table II). It shows significant differences between cognitively-preserved and demented groups concerning the PCG obtained from averaged or middle-slice data. There is a clear separability between the two groups so we suppose that PCG may be used as a predictor of dementia.

Second, to study the predictive potential of PCG, we built ML classification models to prognosticate the diagnosis. The model’s outcome quality was assessed using ROC AUC metric. ROC AUCs, sensitivity, and specificity of the prediction displayed in Table III. Figure 4 illustrates the performance of the NN-based model to predict the diagnosis (HC vs AD) from PCG values predicted from averaged brain images.

**Table III:** Performance of the NN-based classification model to prognosticate the diagnosis out of PCG values

| Cognitive test | Averaged brain |  |  | Middle slice |  |  |
| --- | --- | --- | --- | --- | --- | --- |
|  | Sens. | Spec. | AUC | Sens. | Spec. | AUC |
| ADAS-cog | 0.97 | 0.94 | **0.9871** | 0.97 | 0.94 | 0.9850 |
| MMSE | 0.93 | 0.91 | 0.9780 | 0.95 | 0.94 | **0.9870** |
| RAVLT | 0.85 | 0.78 | 0.8981 | 0.84 | 0.81 | **0.9010** |
| TMT (part B) | 0.90 | 0.73 | **0.8482** | 0.92 | 0.69 | 0.8473 |
| DSST | 0.84 | 0.66 | **0.8292** | 0.82 | 0.64 | 0.7960 |

**Figure 4:**
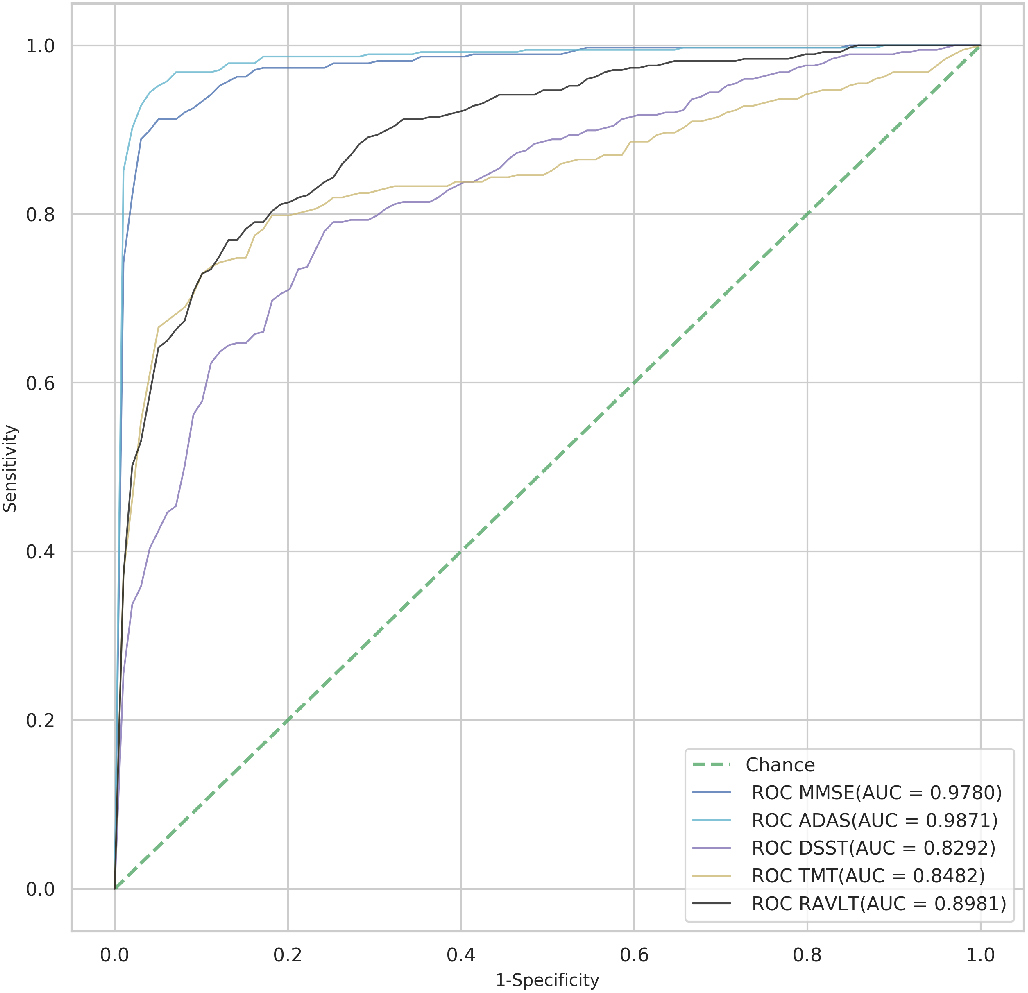
The performance of NN to predict diagnosis from PCG values for cognitive test results. PCG values were obtained from averaged skullstripped brain images.

Cognitive tests used in this study were ranked in descending order for ROC AUC as follows ADAS-cog, MMSE, RAVLT, TMT (part B), DSST. The top listed tests concerning the predictive accuracy on healthy subjects are ADAS-cog and MMSE. These tests are two primary cognitive assessments required in all recent Food and Drug Administration(FDA) clinical drug trials for Alzheimer’s disease in USA [29]. The model’s performance for MMSE and ADAS-cog is comparable to state-of-the-art diagnosis (HC vs AD) classification accuracy based on MRI images [12], [30].

## IV. Conclusions

The comparison of healthy subjects with the demented ones shows significant differences between cohorts concerning cognitive test results. The prediction of the cognitive status similarly to its assessment may reflect the risk of developing dementia.

CNN model can serve as a subtle instrument for the prediction of cognitive decline. The preprocessed T1-weighted MRI images converted to two-dimensional data are a reliable predictor for the level of cognitive impairment.

A single diagnostic test cannot determine if a person has an early stage of dementia. Therefore, physicians have to rely on a set of cognitive tasks commonly named batteries. Ranking the tests with respect to their importance in decision making is crucial for the correct diagnosis. From our data, the potential power of the cognitive tests to predict dementia ranges from high to low in the following order: ADAS-cog, MMSE, RAVLT, TMT (part B), DSST.

The predicted cognitive gap can accurately segregate healthy subjects from demented ones. Therefore, it may serve as a screening tool for dementia.

## Data Availability

Data used in preparation of this article were obtained from the Alzheimer's Disease Neuroimaging Initiative (ADNI) database (adni.loni.usc.edu). As such, the investigators within the ADNI contributed to the design and implementation of ADNI and/or provided data but did not participate in analysis or writing of this report. A complete listing of ADNI investigators can be found at: http://adni.loni.usc.edu/wp-content/uploads/how_to_apply/ADNI_Acknowledgement_List.pdf
All ADNI data are shared without embargo through the https://ida.loni.usc.edu/login.jsp(IDA). Interested scientists may obtain access to ADNI imaging, clinical, genomic, and biomarker data for the purposes of scientific investigation, teaching, or planning clinical research studies. Access is contingent on adherence to the ADNI Data Use Agreement and the publications' policies outlined in the documents listed here http://adni.loni.usc.edu/data-samples/access-data/.

https://ida.loni.usc.edu/login.jsp

## Acknowledgement

The authors would like to acknowledge the College of Information Technology; United Arab Emirates University for offering GPU-based computational facilities such as a supercomputer DGX-1.

